# The Transformative Potential of Large Language Models in Mining Electronic Health Records Data

**DOI:** 10.1101/2024.03.07.24303588

**Authors:** Amadeo Wals Zurita, Héctor Miras del Rio, Nerea Ugarte Ruiz de Aguirre, Cristina Nebrera Navarro, María Rubio Jiménez, David Muñoz Carmona, Carlos Míguez Sánchez

**Affiliations:** Servicio de Oncología Radioterápica. Hospital Universitario Virgen Macarena. Sevilla. España; Servicio de Radiofísica Hospitalaria. Hospital Universitario Virgen Macarena. Sevilla. España

## Abstract

**Introduction:** In this study, we evaluate the accuracy, efficiency, and cost-effectiveness of Large Language Models (LLMs) in extracting and structuring information from free-text clinical reports, particularly in identifying and classifying patient comorbidities within oncology electronic health records. We specifically compare the performance of gpt-3.5-turbo-1106 and gpt-4-1106-preview models against that of specialized human evaluators.

**Methods:** We implemented a script using the OpenAI API to extract structured information in JSON format from comorbidities reported in 250 personal history reports. These reports were manually reviewed in batches of 50 by five specialists in radiation oncology. We compared the results using metrics such as Sensitivity, Specificity, Precision, Accuracy, F-value, Kappa index, and the McNemar test, in addition to examining the common causes of errors in both humans and GPT models.

**Results:** The GPT-3.5 model exhibited slightly lower performance compared to physicians across all metrics, though the differences were not statistically significant (McNemar’s test p = 0.79). GPT-4 demonstrated clear superiority in several key metrics (McNemar’s test p < 0.001). Notably, it achieved a sensitivity of 96.8%, compared to 88.2% for GPT-3.5 and 88.8% for physicians. However, physicians marginally outperformed GPT-4 in precision (97.7% vs. 96.8%). GPT-4 showed greater consistency, replicating the exact same results in 76% of the reports across 10 repeated analyses, compared to 59% for GPT-3.5, indicating more stable and reliable performance. Physicians were more likely to miss explicit comorbidities, while the GPT models more frequently inferred non-explicit comorbidities, sometimes correctly, though this also resulted in more false positives.

**Conclusion:** This study demonstrates that, with well-designed prompts, the LLMs examined can match or even surpass medical specialists in extracting information from complex clinical reports. Their superior efficiency in time and costs, along with easy integration with databases, makes them a valuable tool for large-scale data mining and real-world evidence generation.

## Introduction

Real-world data (RWD) holds immense potential for advancing healthcare by providing a comprehensive view of patient health, disease progression, and treatment outcomes [1]. However, RWD presents significant challenges due to its diverse sources and formats, such as electronic health records, medical imaging, and laboratory results, each with different standards and terminologies. Much of this data is unstructured, like free-text clinical notes, which are difficult to process and analyze. Additionally, missing information is common, leading to gaps that hinder accurate analysis. Advanced methodologies and technologies are needed to effectively extract, standardize, and analyze RWD, ensuring its potential to improve healthcare outcomes is fully realized.

Extracting information from clinical texts has traditionally relied on manual methods, where trained healthcare professionals review and annotate clinical notes to identify relevant information such as diagnoses, treatments, and patient outcomes. This manual process is not only time-consuming and labor-intensive but also prone to human error, leading to inconsistencies and inaccuracies. Additionally, statistical and rule-based approaches have been employed, which depend on predefined patterns and keywords to extract information. However, these methods often fall short in handling the complexity and variability inherent in natural language, resulting in incomplete or inaccurate data extraction.

The rise of artificial intelligence, driven by advances in computing power, has propelled the development of Natural Language Processing (NLP). NLP algorithms can automatically structure information from unstructured clinical texts, facilitating analysis and integration with other clinical data [2], [3], [4], [5]. Earlier NLP systems often relied on rule-based systems and simpler machine learning models, implying limitations such as the need for extensive customization, deep computer science knowledge, significant computational resources, and large volumes of high-quality labeled data. These challenges hinder their widespread adoption and optimal performance across different applications.

Transformer models, a deep learning architecture introduced in the paper “Attention is All You Need” by Vaswani et al. in 2017 [6], have revolutionized the field of NLP, establishing themselves as the foundation upon which modern Large Language Models (LLMs) have been developed. LLMs, such as OpenAI’s Generative Pre-Trained Transformers (GPT), are models trained on vast amounts of text to learn complex linguistic patterns. This enables them to generate text, understand context, perform translations, and carry out other tasks with unprecedented accuracy and fluency. Thanks to this capability, users can interact with these models, instructing them to tackle various problems without the need for additional training.

The GPT-3 model, released in 2020, and its successor, GPT-4 [7], introduced in 2023, represent significant advancements in the ability to understand and generate coherent text. The progression from GPT-3 through GPT-3.5 to GPT-4 marks a significant evolution in OpenAI’s language model capabilities. GPT-4 offers enhanced understanding and generation of text due to its larger training dataset and more refined architecture, resulting in responses that are more accurate, contextually aware, and nuanced compared to its predecessors. This latest version also demonstrates improved performance on a broader array of tasks, including complex reasoning and problem-solving. Additionally, it is multi-modal, capable of processing not only text but also images and audio. However, it is important to note that these models are not specifically designed for medical diagnostic purposes.

Currently, there are numerous LLMs available, such as LLaMA, Mistral, Claude, or BioBERT. However, in the medical field, the ChatGPT models have been the most extensively studied [8], demonstrating strong capabilities in various applications, including interpreting clinical guidelines and enhancing evidence-based medicine [9], or table summarization in clinical study reports [10]. Despite their potential, concerns about the applicability of these general-purpose models in the medical domain persist [11], [12], particularly due to their lack of transparency in training data, which remains largely unknown. Therefore, it is essential to evaluate their performance for each specific application.

In the context of extracting and structuring information from free-text clinical reports, studies have shown promising results with OpenAI models. For instance, Matthias A. Fink et al. [13] demonstrated the effectiveness of these models in extracting data from computed tomography reports related to lung cancer, where they outperformed traditional NLP models in classifying disease progression.

Focusing on the significance of appropriate instructions (prompts), researchers such as Hyeon Seok Choi et al [14] highlighted that the gpt-3.5-turbo model exhibited an accuracy rate of 87.7% in extracting information from pathology and ultrasound reports of breast cancer patients. Additionally, the LLM methods demonstrated superior efficiency in terms of time and costs compared to manual approaches.

In 2018, the Department of Radiation Oncology at HUVM initiated the implementation of the Mosaiq system, transitioning towards a paperless workflow and centralizing all radiation therapy treatment data within the application. As detailed by Bertolet *et al* [15], this data was automatically exported to JSON files via Word documents and VBA code. Figure 1 depicts a diagram illustrating the flow and organization of the described data.

**Figure 1:**
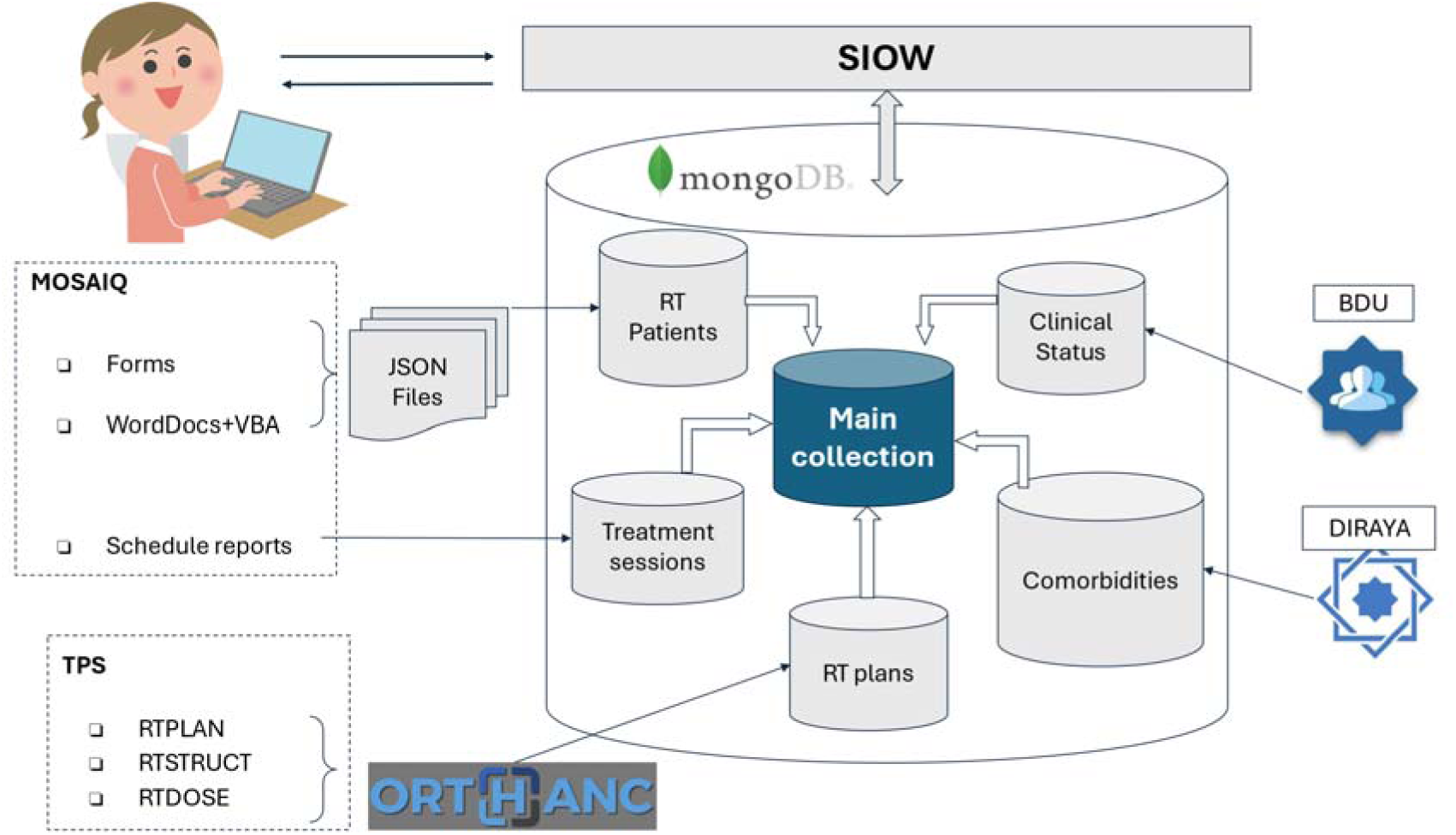
Representative diagram of the Web Oncological Information System (SIOW). It illustrates the integration of data from MOSAIQ and TPS into the MongoDB database, and its subsequent management through SIOW, including the collection of administrative data from the Users Data Base (BDU) and clinical data from the EHR system DIRAYA.

Motivated by the capabilities of LLMs, we aimed to investigate their potential application in extracting and structuring information from clinical reports. Our overarching objective is to integrate LLM-based tools into our information system, enhancing the richness of our real-world datasets. Specifically, in this study, we assess the capability of the GPT-3.5 turbo and GPT-4 models as tools for data mining applied to the identification and classification of comorbidities and relevant lifestyle risk factors in oncological texts. We compare their performance against that of specialized human evaluators to gauge their efficacy and suitability for clinical use.

## Methods

### OpenAI models

The application programming interface (API) of OpenAI [16] allows interaction with their advanced LLMs, facilitating various language processing tasks such as generating automatic textual responses, conducting sentiment analysis, and summarizing texts. In our study, we leverage the *chat completions API* function of the API to extract structured information from unstructured clinical reports.

OpenAI offers a comprehensive library of natural language processing models. Each model features unique characteristics in terms of size, language comprehension ability, speed, and cost. In our study, we have employed two models from the library: ***gpt-3.5-turbo-1106*** and ***gpt-4-1106-preview***, with the latter being the most advanced model available at the time the study was conducted. While the GPT-3.5 model is a faster and more economical option for general tasks, GPT-4 stands out for its higher accuracy, contextual understanding, and ability to handle more complex and specific applications.

For this study, we utilized clinical reports in Spanish, exclusively interacting with OpenAI’s LLMs in this language. Although LLMs typically exhibit superior performance in English [17], owing to the predominance of this language in training data, recent comparisons indicate notable effectiveness in other languages, including Spanish. The GPT-4 technical report [7] highlights this multilingual capability, demonstrating that performance in Spanish closely approaches that of English, with a minimal difference of only 1.5 percentage points in the MMLU evaluation [18].

### Prompt generation

To interact with the LLM models, we first created a prompt that will guide the model through the specific task. The context provided to the model establishes a scenario in which it is asked to assume the role of a specialist in radiation oncology. This setting serves as a reference framework, enabling the model to adopt the appropriate perspective and apply its natural language understanding capabilities in a manner consistent with the medical domain.

Our request is a direct instruction to the model, directing it to process the text of the provided clinical report and return the relevant information in a structured format. Specifically, the model is instructed to utilize the clinical report provided at the end of the prompt to complete a predefined dictionary in JSON format. This dictionary contains keys related to comorbidities and lifestyle risk factors. The model is tasked with updating the values of these keys with “YES” or “NO” as appropriate. For individuals who are ex-smokers, the model should use “EX” instead. Additionally, the model must identify and add any other relevant comorbidities not classifiable under the provided categories, assigning them to the “Other” key.

This is the prompt generated for the task:

- **Context**: “Act as a specialist in radiation oncology.”
- **Request**: “Use the clinical report provided at the end of this prompt to return in JSON format the dictionary […] with the values ’YES’ or ’NO’. For the ’Smoker’ field: ’YES’ if they smoke, ’NO’ if they have never smoked, ’EX’ if they are an ex-smoker. For the ’Other’ field, return a list of comorbidities found that cannot be classified in any of the categories of the keys of the provided dictionary, or empty if there are no other comorbidities. Return only the dictionary with the updated values, DO NOT ADD OR MODIFY KEYS. Clinical report: [text of the clinical report]”

The dictionary mentioned in the request is structured with keys labeling the specific comorbidities and lifestyle risk factors we seek to identify. These comorbidities, along with their potential values, are outlined in Table 1.

**Table 1:** List of the labels, possible values, and description of the comorbidities and lifestyle risk factors considered in this study.

| Label | Values | Description |
| --- | --- | --- |
| Diabetes | YES/NO | Elevated blood glucose levels |
| HBP | YES/NO | High Blood Pressure |
| Smoker | YES/NO/EX | Smoking habit. |
| Dyslipidemia | YES/NO | Lipid metabolism disorder |
| Liver Disease | YES/NO | Liver Disease |
| COPD | YES/NO | Chronic Obstructive Pulmonary Disease |
| Depression | YES/NO | Mood disorder |
| Kidney Disease | YES/NO | Kidney Disease |
| Fentanyl | YES/NO | Use of WHO step 3 analgesics. (Opioids) |
| Heart Disease | YES/NO | Heart Disease |
| Hyperthyroidism | YES/NO | Thyroid disease with increased thyroxine |
| Hypothyroidism | YES/NO | Thyroid disease with decreased thyroxine |
| Dependent | YES/NO | Patient in need of continuous care |
| Other | Text list | Other past comorbidities detected not listed above. |

During a postprocessing phase, we divided the category labeled as “*smoker*” into two distinct categories: “*smoker*” (representing current smokers) and “*ex-smoker*”. This division was implemented to ease the subsequent analysis of the results.

It’s important to highlight that the prompt does not provide context or additional instructions regarding how the specified comorbidities of interest should be interpreted.

The development of this prompt was achieved through an iterative process applied to a group of 50 reports that were specifically reserved for this purpose. The methodology included the following steps:

1. **Prompt Definition:** Establishing the parameters and structure of the prompt to guide the model’s responses.
2. **Information Extraction:** The developed prompt was applied to 50 reports using the *gpt-4-1106-preview* model.
3. **Verification of Structure:** It was ensured that the model’s responses adhered to the requested structure, with previous steps being repeated in case of deviations.
4. **Accuracy Evaluation:** A specialist physician (AW) verified the accuracy of the model’s responses. This process was repeated until the accuracy met or exceeded that of a manual analysis performed by the same physician.

### Python Script

The Python script developed utilizes the OpenAI API to automatically structure textual clinical information. All the code developed for this work is publicly available in a GitHub repository: https://github.com/RFMacarena/openaiAPIscript_forsharing.

### Clinical Report Acquisition Procedure

The clinical reports for our study were provided by the hospital’s Innovation & Data Analysis department. These reports were delivered in an Excel spreadsheet format, organized into two essential columns: one containing the clinical history number of each patient and another with the text of the medical personal history report. The department responsible for data collection undertook a process of anonymization and randomization of the reports to ensure an unbiased selection.

### Sample Selection Criteria

For estimating the sample size, we relied on the proportion of comorbidities (80%) obtained from a prior analysis of a dataset of 5257 personal history reports from patients treated in our service between May 2018 and October 2022.

The comorbidities selected for the study were chosen based on prior knowledge of prevalences in the general population and those presented by our patients according to the aforementioned analysis. We also considered those that could most significantly impact the clinical outcome of oncological treatments.

With these considerations, we conducted a preliminary calculation that established the need to include 250 clinical reports (see below in the statistical analysis section). Based on this calculation, we selected the first 250 patients from the provided list who had a non-empty personal history report. Before proceeding with the analysis, we verified that our script was capable of correctly interpreting an empty report as equivalent to the absence of comorbidities, thereby avoiding biases in the study results.

### Ethical Considerations and Data Protection

The text processed by the selected LLMs is strictly confined to personal history reports. These reports were stripped of any information that could lead to patient identification, ensuring confidentiality and anonymity. The model’s interpretation of the texts focuses solely on identifying and structuring data relevant to the study, without compromising individual privacy.

The study’s design, synthesized in Figure 2, and methodology were previously communicated to and reviewed by the hospital’s ethics committee. The research received the necessary approval, confirming that it adheres to the ethical standards required for patient data research.

**Figure 2:**
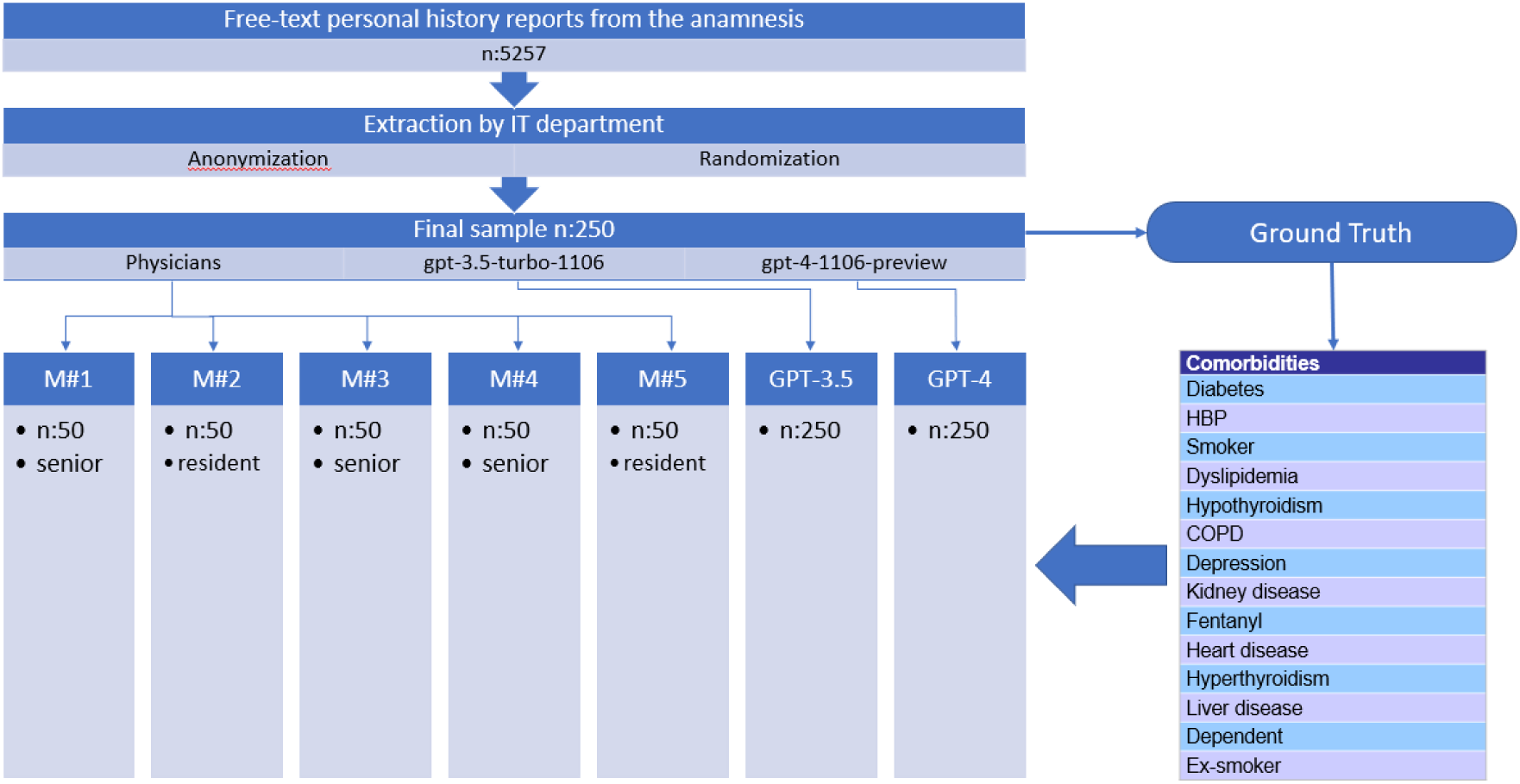
Flowchart of the study design.

This retrospective study adheres to the guidelines outlined in the *seventeenth additional provision, specifically Health Data Processing, Section d) of the Organic Law 3/2018, dated December 5, on Personal Data Protection and Guarantee of Digital Rights*. This law governs the use of pseudo-anonymized personal data for health research purposes. The study was granted an exemption from requiring informed consent due to its exclusive use of non-identifiable data.

On January 18, 2024, the Ethics Committee of the University Hospitals Virgen Macarena and Virgen del Rocío issued a favorable opinion for our study, under the reference EC_IA_V1 (Version 1-Dec-2023).

### Data Extraction

For the manual data extraction, the 250 patient clinical reports were divided into five groups, each consisting of 50 reports. These groups were randomly assigned to five physicians, including three specialists in radiation oncology with more than 15 years of experience and two medical residents in the same specialty, one in their first year and the other in their fourth year.

To ensure uniform and accurate data collection, the physicians were provided with a specially designed template for this task. The template features a table where the first column contains the full texts of the clinical reports. The subsequent columns of the table are labeled with the comorbidities of interest. The cells corresponding to each comorbidity only allow the selection of predefined values, as stipulated in Table 1. This restriction ensures consistent annotation and reduces the possibility of errors or variations in the entries.

For the automatic analysis, the 250 clinical reports in the sample were analyzed using our script with the *gpt-3.5-turbo-1106* and *gpt-4-1106-preview* models. To maintain a consistent structure in the study, these reports were organized into the same five groups of 50 reports that were assigned to the physicians. The results were recorded in a document that mirrored the structure of the template used in the manual extraction. This uniformity in documentation facilitates a direct comparison of results between manual and automatic extraction methods.

### Establishing the Ground Truth

To assess the comparative accuracy and effectiveness of the LLMs used in this study against the evaluations performed by physicians, it is crucial to establish a reference dataset containing the ground truth. To construct this reference dataset, we first compared the results obtained from the physicians and the *gpt-4-1106-preview* model across all 250 reports, identifying and recording any discrepancies between the two sources. The radiation oncologist expert AW, with more than 30 years of experience, reviewed several times the whole set of reports, with a particular focus on these discrepancies. For each report where discrepancies in the results were found, physician AW assessed both responses (from the physician and the AI) and determined which one was correct.

It’s important to note that the ground truth in this study is based solely on the information explicitly reported in the clinical texts. This means that some patients may have unreported comorbidities, or conversely, conditions may be mentioned that are not actually present. This limitation reflects a common challenge when working with real-world data. However, for the purposes of this study, these potential discrepancies are irrelevant, as our primary focus is on evaluating the models’ ability to accurately interpret and extract information from the provided texts.

### Assessing Reproducibility in Results

The non-deterministic nature of LLMs, such as GPT-3.5 and GPT-4, means they can generate different responses to identical requests [7]. This phenomenon, coupled with the potential for periodic retraining of the models, significantly impacts the reproducibility of results. Therefore, it is crucial to consider the need for rigorous quality control for algorithms that employ LLMs, especially to assess the impact of any changes in the models.

A well-defined and explicit prompt can increase the reproducibility of responses [14]. However, variability remains a possibility, particularly in situations where the information is ambiguous, or the prompt is not clear or specific enough.

To measure the consistency of our automatic extraction method, we repeated the analysis of the 250 clinical reports 10 times over 10 consecutive days. This approach allows us to observe the stability of the model responses to the same input.

### Statistical Analysis

To ensure the statistical validity of the study, a significance level of 5% (alpha error) and a power of 80% (beta error of 20%) were established. Additionally, a 5% error margin was applied for 95% confidence intervals. With these considerations in mind, it was determined that the sample size (n) should include 245 patient records. To adjust the sample to a practical number, it was rounded up, resulting in a final sample size of 250.

For a comprehensive analysis, we consolidated the results from the 250 reports into a single category named “Physicians,” representing the aggregated findings of the five doctors involved in the study. Subsequently, we compared this category and the results from the GPT-3.5 and GPT-4 models with the reference dataset, considered as the ground truth. In this process, a confusion matrix was created for each report and comorbidity, from which several key statistical estimators were derived.

To assess the agreement, we employed the Kappa index. McNemar’s test was used to determine if there were significant differences in the proportions of discordance between the classifications. We chose the F-score as a measure of balance between precision and sensitivity, which is crucial in a classification model. The calculated metrics are presented in Table 2.

**Table 2:** Metrics used in the study with their descriptions.

| Metric | Description |
| --- | --- |
| TP | True Positives |
| TN | True Negatives |
| FP | False Positives |
| FN | False Negatives |
| Sensitivity | $TP / (TP+FN)$ |
| Specificity | $TN / (FP+TN)$ |
| Precision | $TP / (TP+FP)$ |
| Prevalence | $(TP+FN) / (TP+TN+FP+FN)$ |
| Accuracy | $(TP+TN) / (TP+TN+FP+FN)$ |
| Kappa | $(P_{obs}-P_{esp}) / (1-P_{esp})$ |
| F-score | $(2 * Precision * Sensibility) / (Precision + Sensibility)$ |
| McNemar | Exact-P-value from McNemar test (binomial distribution) |

For some of these metrics, we calculated their confidence interval using the bootstrapping method [19]. This approach starts from the frequencies of True Positives (TP), True Negatives (TN), False Positives (FP), and False Negatives (FN) to generate 1,000 resamples. With these resamples, we recalculated the metrics to obtain a distribution that allows us to calculate the 95% confidence interval.

Additionally, a detailed analysis was conducted on the groups of 50 reports assigned to each physician. This analysis focused on measuring the variability in evaluations among different physicians. For each patient and comorbidity, Cohen’s kappa index was calculated in comparison with the ground truth for the results of each physician.

The reproducibility of the GPT-3.5 and GPT-4 models was assessed by quantifying the number of different responses for each patient and comorbidity across the 10 repeated analyses conducted on successive days.

### Analysis of Discrepant Results

A detailed analysis of discrepancies between the evaluators’ results and the established Ground Truth was conducted by the same physician who defined the reference dataset. This analysis covered each report with discrepancies in the identification of comorbidities, identifying the probable causes of each deviation.

Discrepancies were classified according to the nature of the detected errors:

- **Differences in criteria:** Variations in the interpretation of the relevance of reported pathologies.
- **Incorrect interpretation:** Misunderstandings caused by confusing wording.
- **Incorrect inference:** Erroneous deductions when the comorbidity is not explicitly mentioned.
- **Ambiguous text:** Textual ambiguity that allows for multiple interpretations.
- **Error/Hallucination:** Unjustified errors, attributed to human distractions or AI hallucinations.
- **Error in Ground Truth:** Corrections made upon review that validate the evaluator’s interpretation.
- **Explicit omission:** Overlooking direct mentions of comorbidities.
- **Omission by context:** Failure to notice comorbidities deducible from the context or medication.
- **Unrecognized acronyms:** Inability to interpret specific medical acronyms.

## Results

### Cost and time analysis

Table 3 details the cost and total time invested in analyzing the 250 reports using the GPT-3.5 and GPT-4 models. Given that both the models and their associated costs can fluctuate over time, it is important to note that the reported results are specific to the usage period from January to February 2024. It is noted that GPT-4, being a larger and more complex LLM compared to GPT-3.5, incurs longer processing times and a cost approximately 10 times higher. Extrapolating the costs to the entire set of 7,500 patients currently registered in our database, processing with GPT-4 would require about 24 hours and would cost approximately 76 dollars. On the other hand, using GPT-3.5 would reduce the processing time to about 9 hours, with a significantly lower cost of around 7 dollars.

**Table 3:**
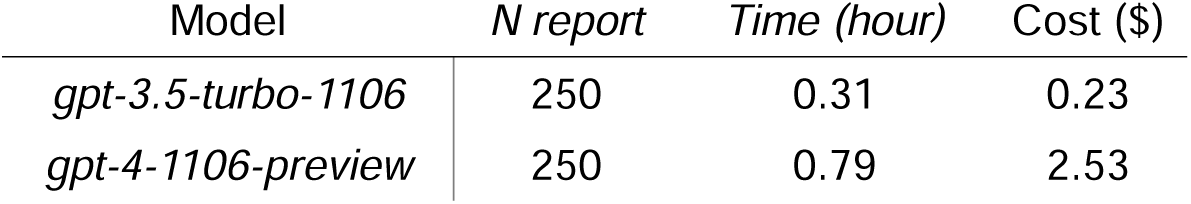
Execution times and costs in dollars for the analysis of the 250 reports with each of the models used (Usage period of the models: between January and February 2024).

| Model | *N report* | *Time (hour)* | Cost (\$) |
| --- | --- | --- | --- |
| *gpt-3.5-turbo-1106* | 250 | 0.31 | 0.23 |
| *gpt-4-1106-preview* | 250 | 0.79 | 2.53 |

### Prevalences

The analysis of our Ground Truth sample reveals a wide range of prevalences in comorbidities and lifestyle risk factors among oncological patients. These are detailed in Table 4, where both the number of cases and the prevalence for each comorbidity are reported. The most common conditions include high blood pressure and dyslipidemia, present in almost half and a third of the cases, respectively. On the other hand, conditions like hyperthyroidism and liver disease show relatively low prevalence. Categories related to smoking are also highly frequent, accounting for almost 50% of the cases. Interestingly, the proportion of ex-smokers significantly exceeds that of current smokers.

**Table 4:** Number of reports, out of the total 250 in the sample, that indicate each comorbidity and the corresponding prevalence.

| Condition | #Cases | Prevalence |
| --- | --- | --- |
| Diabetes | 64 | 25.6% |
| HBP | 116 | 46.4% |
| Smoker | 37 | 14.8% |
| Dyslipidemia | 77 | 30.8% |
| Hypothyroidism | 21 | 8.4% |
| COPD | 17 | 6.8% |
| Depression | 25 | 10.0% |
| Kidney disease | 39 | 15.6% |
| Fentanyl | 19 | 7.6% |
| Heart disease | 43 | 17.2% |
| Hyperthyroidism | 1 | 0.4% |
| Liver disease | 13 | 5.2% |
| Dependent | 12 | 4.8% |
| Ex-smoker | 85 | 34.0% |

### Evaluation metrics

Table 5 displays the values of true positives, false positives, true negatives, and false negatives, detailed by comorbidity, derived from the comparison with the Ground Truth dataset.

**Table 5:** Tables displaying the results for true positives (TP), true negatives (TN), false positives (FP), and false negatives (FN) for each comorbidity, obtained by each of the evaluators (Physicians, GPT-3.5, and GPT-4).

|  | Physicians |  |  |  | GPT-3.5 |  |  |  | GPT-4 |  |  |  |
| --- | --- | --- | --- | --- | --- | --- | --- | --- | --- | --- | --- | --- |
|  | TP | TN | FP | FN | TP | TN | FP | FN | TP | TN | FP | FN |
| Diabetes | 63 | 185 | 1 | 1 | 54 | 186 | 0 | 10 | 63 | 186 | 0 | 1 |
| HBP | 110 | 133 | 1 | 6 | 113 | 132 | 2 | 3 | 114 | 133 | 1 | 2 |
| Smoker | 37 | 209 | 4 | 0 | 36 | 212 | 1 | 1 | 36 | 213 | 0 | 1 |
| Dyslipidemia | 67 | 173 | 0 | 10 | 67 | 172 | 1 | 10 | 74 | 173 | 0 | 3 |
| Hypothyroidism | 19 | 229 | 0 | 2 | 19 | 229 | 0 | 2 | 20 | 227 | 2 | 1 |
| COPD | 16 | 233 | 0 | 1 | 15 | 233 | 0 | 2 | 16 | 231 | 2 | 1 |
| Depression | 25 | 224 | 1 | 0 | 21 | 220 | 5 | 4 | 22 | 223 | 2 | 3 |
| Kidney disease | 15 | 211 | 0 | 24 | 21 | 211 | 0 | 18 | 38 | 208 | 3 | 1 |
| Fentanyl | 18 | 231 | 0 | 1 | 18 | 230 | 1 | 1 | 19 | 230 | 1 | 0 |
| Heart disease | 38 | 207 | 0 | 5 | 30 | 205 | 2 | 13 | 40 | 205 | 2 | 3 |
| Hyperthyroidism | 0 | 249 | 0 | 1 | 0 | 249 | 0 | 1 | 1 | 249 | 0 | 0 |
| Liver disease | 9 | 236 | 1 | 4 | 12 | 234 | 3 | 1 | 13 | 234 | 3 | 0 |
| Dependent | 12 | 234 | 4 | 0 | 11 | 238 | 0 | 1 | 10 | 238 | 0 | 2 |
| Ex-smoker | 76 | 165 | 0 | 9 | 85 | 161 | 4 | 0 | 85 | 163 | 2 | 0 |
| *Total* | *505* | *2919* | *12* | *64* | *502* | *2912* | *19* | *67* | *551* | *2913* | *18* | *18* |

Figure 3 illustrates the performance of the physicians, GPT-3.5, and GPT-4 classifiers, broken down by comorbidity, across various metrics. The “Total” category, which consolidates the results for all studied comorbidities, enables direct comparison between the three evaluators on each assessed metric:

- **Sensitivity:** The GPT-4 model (96.8%) outperforms both GPT-3.5 (88.2%) and the physicians (88.8%) in most categories, showing notable effectiveness in detecting comorbidities. Although GPT-3.5 presents slightly lower results than the physicians, the difference is not statistically significant, as indicated by the overlap of the 95% confidence intervals shown in Figure 3.
- **Specificity:** All evaluators achieve high specificity values, which is expected given the low prevalences of the studied comorbidities and the relative ease of identifying the absence of a comorbidity in texts. The physicians (99.6%) excel in this metric, often achieving perfection, while both models (99.4%) score slightly lower due to a higher rate of false positives.
- **Precision:** The physicians get the highest score (97.7% vs 96.4% and 96.8%) assessing the proportion of correct positive identifications, possibly also influenced due to the models generating a higher number of false positives.
- **F-Score:** Representing the harmonic mean between precision and sensitivity, the F-Score is particularly relevant in asymmetric samples like in our study. The GPT-4 model achieves the highest score (96.8%) on this indicator, surpassing both GPT-3.5 (92.1%) and the physicians (93.0%).
- **Accuracy (Agreement):** In the proportion of correct identifications, GPT-4 shows superior performance (99.0%), while GPT-3.5 (97.5%) and the physicians (97.8%) achieve similar results.
- **Cohen’s Kappa Index:** This index, measuring agreement adjusted for chance, reveals that GPT-4 reaches the highest scores (0.962), demonstrating greater consistency compared to the ground truth. The GPT-3.5 score of 0.907, while marginally lower, does not significantly differ from the physicians’ score of 0.917.

**Figure 3:**
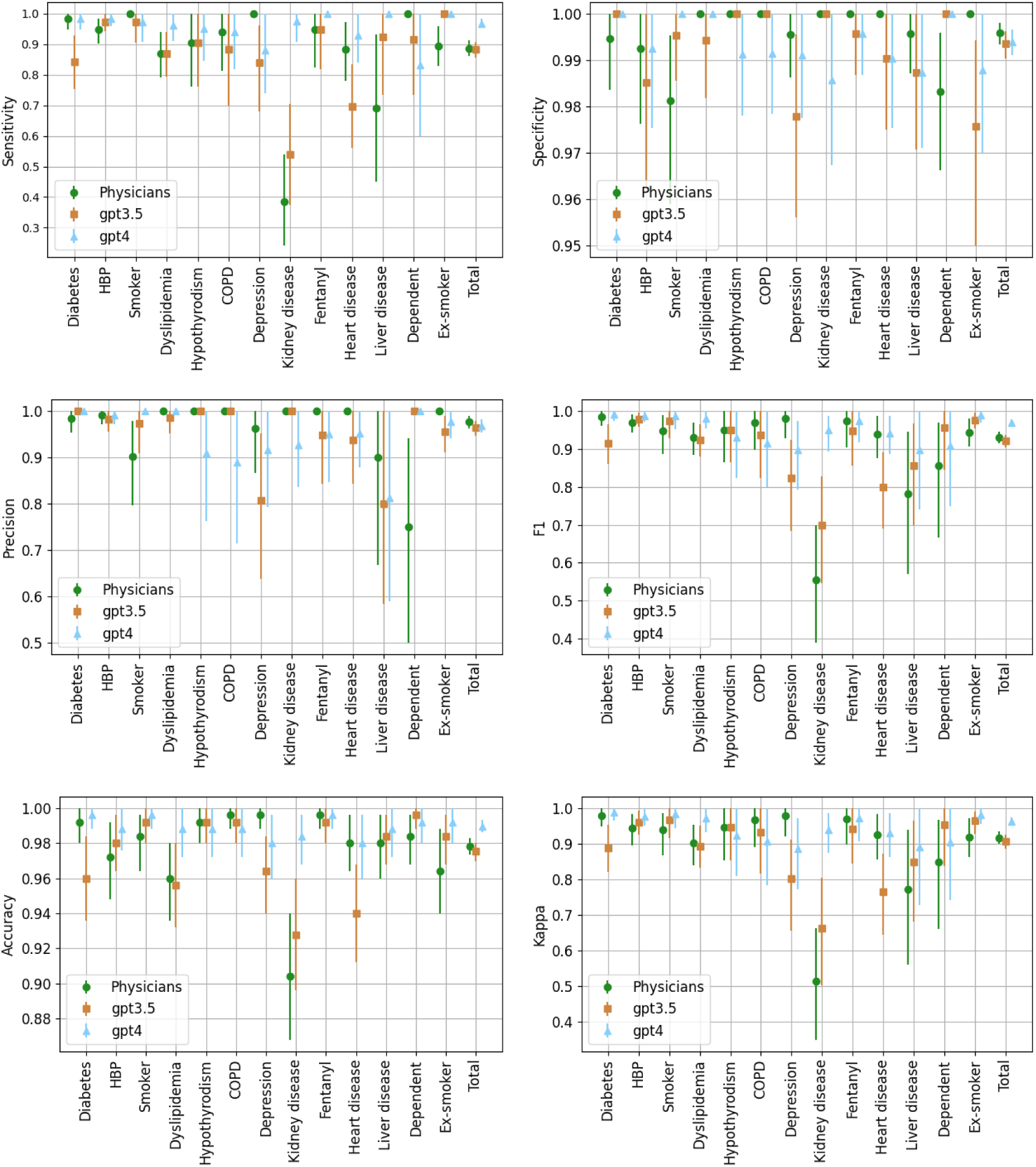
Statistical metrics comparison between three evaluators (Physicians, GPT-3.5, and GPT-4) for individual comorbidities and overall totals. Asymmetric error bars indicate the 95% confidence interval.

The application of McNemar’s test to the “Total” category, comparing Physicians with GPT-3.5 and Physicians with GPT-4, yielded p-values of 0.79 and 10^-6^, respectively. This confirms that the performance differences between the physicians and the GPT-3.5 model are not statistically significant, while the differences between the physicians and GPT-4 are significant.

### Variability among physicians’ performance

Table 6 displays the Cohen’s Kappa index values obtained in the detection of various comorbidities for each of the five physician evaluators. It is important to note that each physician analyzed a different group of 50 reports.

**Table 6:** Concordance values for each comorbidity, calculated using Cohen’s Kappa index for each medical evaluator. The “Total” categories summarize the aggregated concordance across all comorbidities and medical evaluators. A dash indicates that the Kappa index could not be computed because the comorbidity was not present in the corresponding set of reports.

|  | #M1<br>senior | #M2<br>resident | #M3<br>senior | #M4<br>senior | #M5<br>resident | TOTAL<br>#HUM |
| --- | --- | --- | --- | --- | --- | --- |
| Diabetes | 1.00 | 0.95 | 1.00 | 1.00 | 0.95 | 0.98 |
| HBP | 1.00 | 0.96 | 0.83 | 0.96 | 0.96 | 0.94 |
| Smoker | 1.00 | 1.00 | 0.88 | 0.86 | 0.93 | 0.94 |
| Dyslipidemia | 0.91 | 1.00 | 0.75 | 0.77 | 1.00 | 0.90 |
| Hypothyroidism | 0.66 | 1.00 | 0.90 | 1.00 | 1.00 | 0.95 |
| COPD | 1.00 | 1.00 | 1.00 | 0.66 | 1.00 | 0.97 |
| Depression | 0.93 | 1.00 | 1.00 | 1.00 | 1.00 | 0.98 |
| Kidney disease | 0.52 | 0.70 | 0.45 | 0.56 | 0.26 | 0.51 |
| Fentanyl | 1.00 | 1.00 | 0.85 | 1.00 | 1.00 | 0.97 |
| Heart disease | 0.95 | 1.00 | 0.91 | 0.79 | 1.00 | 0.93 |
| Hyperthyroidism | - | - | - | - | 0.00 | 0.00 |
| Liver disease | - | 1.00 | 0.63 | 0.65 | 1.00 | 0.77 |
| Dependent | 0.66 | 0.66 | 0.91 | 0.88 | - | 0.85 |
| Ex-smoker | 0.95 | 1.00 | 0.87 | 0.76 | 1.00 | 0.92 |
| Total | 0.95 | 1.00 | 0.87 | 0.76 | 1.00 | 0.92 |

Overall, there was considerable similarity in the physicians’ responses, except when the comorbidity to be detected was a broader concept, as in the case of “kidney disease” (kappa 0.51) or “liver disease” (kappa 0.77). It’s important to note that no further instructions or explanations were provided beyond finding the comorbidity in the presented text. Therefore, some physicians considered that renal lithiasis was not a relevant “kidney disease” and reserved this category for conditions describing an alteration in renal function (such as chronic renal failure, for example).

Interestingly, the senior physicians scored lower than the medical residents in the overall calculation for the Kappa index.

### Reproducibility of models’ responses

In our reproducibility study, each report was analyzed 10 times by the GPT-3.5 and GPT-4 models. For each comorbidity, we counted the number of different responses generated in these repeated analyses, as well as the total number of variations for each report.

Figure 4 presents a histogram illustrating the number of reports that generated at least the specified number of different responses. This histogram reveals that, in all instances, the GPT-4 model exhibited fewer differences in responses compared to GPT-3.5, suggesting greater consistency and reliability in its results.

**Figure 4:**
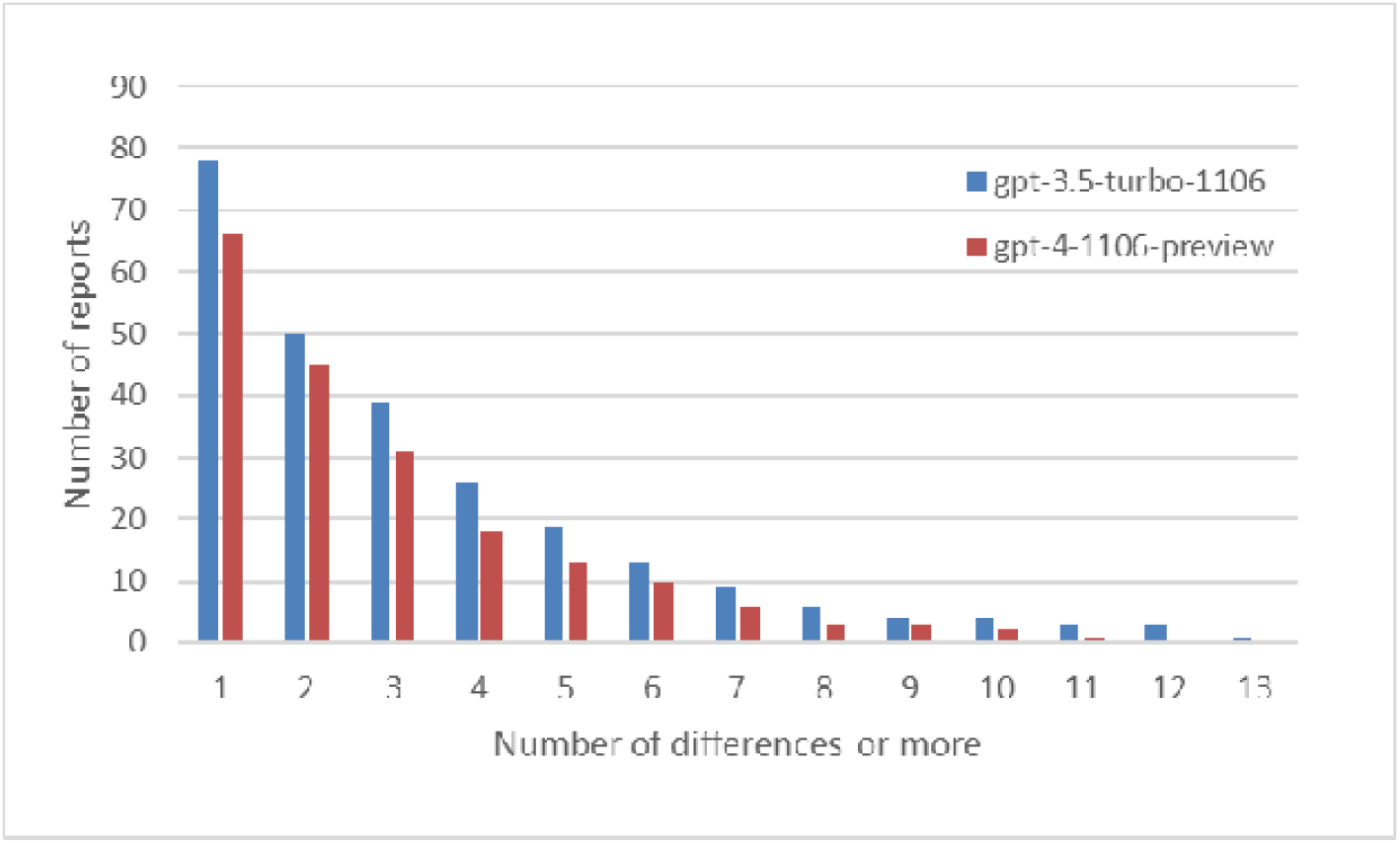
For each model, the number of reports is shown in which at least the number of differences indicated on the x-axis were obtained in the 10 analyses.

Furthermore, it was found that 73.6% of the reports analyzed with GPT-4 reproduced the same result across all comorbidities during the 10 analyses, compared to 59.2% for GPT-3.5. This notable difference in reproducibility underscores the superiority of GPT-4 in maintaining consistency in its responses across multiple executions.

Variability in responses often stems from ambiguous text, where LLMs may assign values inconsistently. For example, a report describing a patient as an “active smoker (1 month since quitting, 1 pack/day since age 14-16)” resulted in GPT-3.5 identifying the patient as a smoker in six out of ten analyses, while GPT-4 made only one error across ten analyses. However, in the same report, regarding the comorbidity of COPD, GPT-4 shows a split: in five instances it identifies it as present and in five as absent. The physician reviewing the results and establishing the ground truth determined the absence of COPD, as it is not explicitly mentioned in the report. Nonetheless, the mention of “mild pulmonary emphysema areas” and the patient’s prolonged smoking history could lead GPT-4 to infer the presence of COPD.

### Discrepancy Analysis

Figures 5 and 6 display the distribution of discrepant results categorized by the causes determined through a detailed manual analysis of the reports.

**Figure 5:**
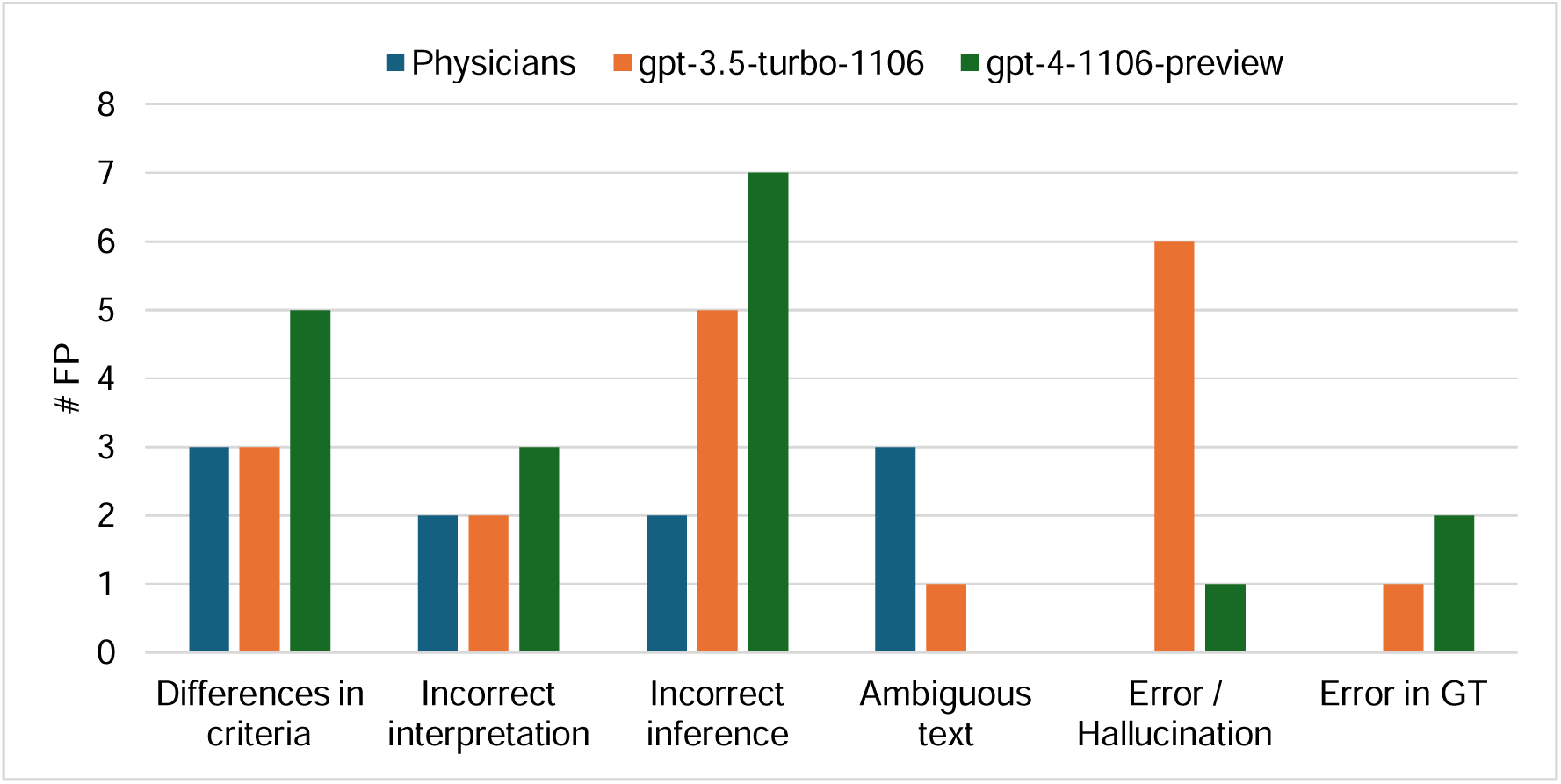
Number of false positive results attributed to each of the considered causes.

**Figure 6:**
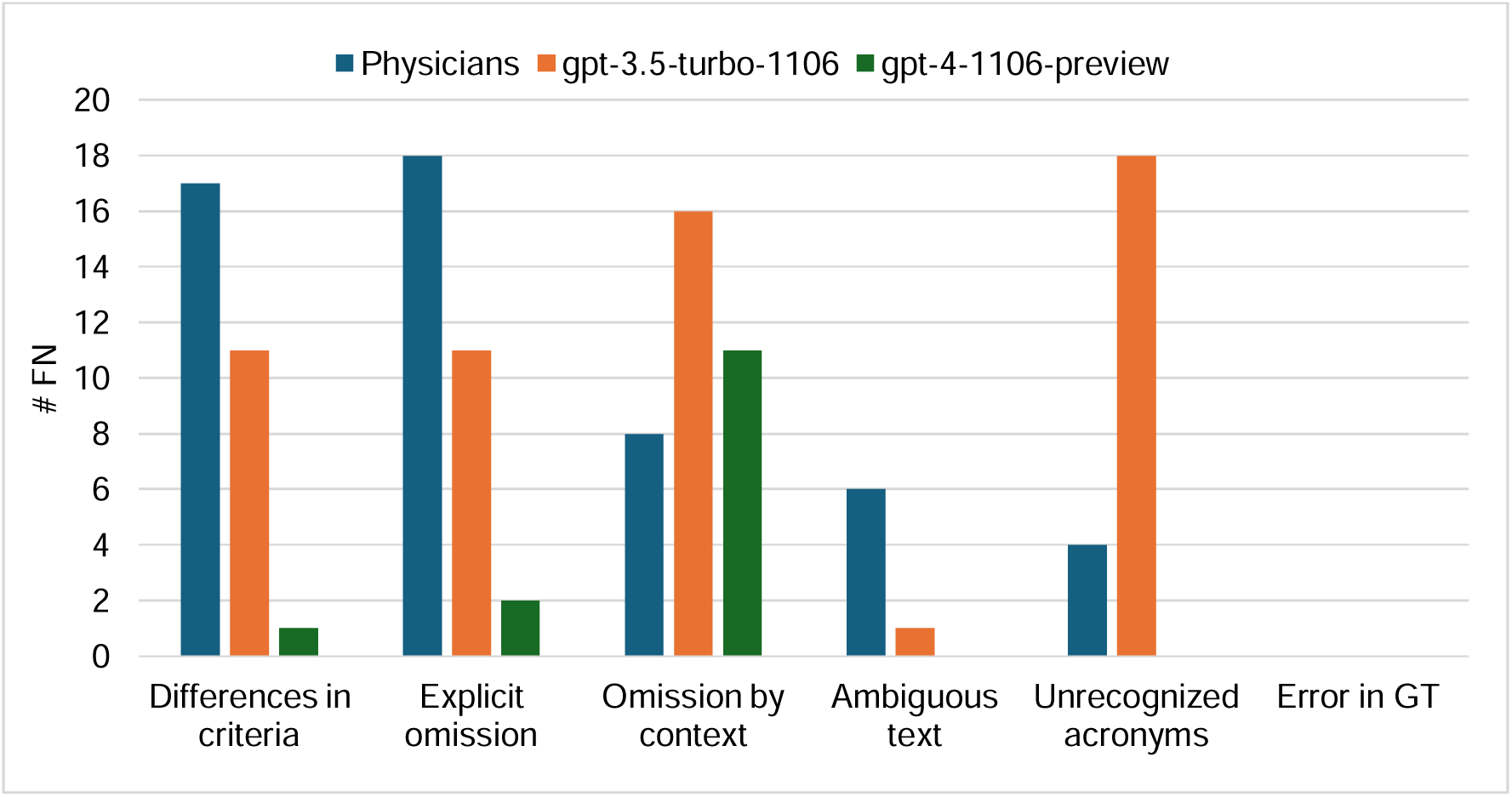
Number of false negative results attributed to each of the considered causes.

A notable discrepancy arose in the “kidney disease” category due to differences in criteria. Some physicians and GPT-3.5 did not deemed certain renal pathologies, such as renal lithiasis, as relevant comorbidities in the context of oncology treatment, unlike GPT-4, which aligned its results more closely with the ground truth.

In analyzing cases interpreted as hallucinations, it was found that this phenomenon occurred exclusively in 1 response from GPT-4 and in 6 from GPT-3.5, particularly in the smoker and ex-smoker categories, possibly due to the use of the label “toxic habits,” even when referring to other habits like alcoholism.

The models, especially GPT-4, tend to infer comorbidities from the context or reported medication more frequently than physicians, who exhibit a more conservative approach. This tendency leads to more false positives by the models, particularly when the medication does not imply the presence of comorbidity.

GPT-3.5 exhibited difficulties in interpreting common medical acronyms such as “DM” for diabetes or “AF” for atrial fibrillation, whereas GPT-4 demonstrated a superior ability to recognize and correctly interpret most of these acronyms.

Interestingly, GPT-4 displayed some false positives when encountering comorbidity labels followed by “:” without additional information, a misinterpretation not common in humans but observed in AI, particularly in GPT-4 more than in GPT-3.5.

Human evaluators showed a greater tendency to overlook comorbidities explicitly reported, likely due to distraction or fatigue.

Only three errors were identified in the determination of the ground truth, underscoring the reliability of the review process.

Finally, we identified a category of discrepancies exclusive to the models, related to structural or formatting errors. This includes situations where the models’ responses do not follow the guidelines specified in the prompt, resulting in outputs that do not meet the expected JSON format or that incorrectly alter and/or introduce comorbidity labels. Given that these incidents were limited, affecting less than 10 cases, it was decided to manually correct these formatting errors for inclusion in the subsequent analysis.

## Discussion

Our study categorizes observers as “Physicians,” “GPT-3.5,” and “GPT-4,” reflecting the synergy between specific models (*gpt-3.5-turbo-1106* and *gpt-4-1106-preview*) and the prompts designed for this research. The effectiveness of GPT models in generating responses is inherently linked to the quality and structure of the prompts [14], [20], [21], indicating that results may vary significantly with prompt redefinition. Similarly, physician performance is influenced not only by clinical competence but also by the clarity of instructions and the quality of the materials provided. Offering more detailed and specific guidelines, along with access to additional sources within the EHRs, could potentially improve the accuracy of their responses.

It is important to emphasize that even if LLMs demonstrate superiority in the specific task of processing large volumes of reports to extract information, this should not be extrapolated to other tasks, such as decision-making. In such cases, these tools should always be used as support tools, requiring ongoing physician oversight and intervention.

Based on the results obtained, we can conclude that the GPT-4 model is notably better at identifying present comorbidities, with fewer false negatives, while physicians exhibit slightly higher precision in their diagnoses, resulting in fewer false positives. The GPT-3.5 model generally performs slightly below the physicians, though the differences found are not statistically significant. These results are consistent with findings from other studies, such as Hoppe et al. [22], which highlight the potential of ChatGPT models to enhance diagnostic accuracy in emergency medical settings. In their study, GPT-4 also outperformed both resident physicians and GPT-3.5 in diagnostic accuracy.

The superior sensitivity of GPT-4 in our study is particularly noteworthy, demonstrating its advanced ability to accurately identify reported comorbidities, even when not directly evident in the text. However, both GPT-3.5 and GPT-4 generate a comparable number of false positives, which is significantly higher than those recorded by physicians. Physicians’ false positives typically result from specific circumstances such as ambiguity in clinical reports, variations in interpretation among professionals, and occasional errors in the template filling process.

In contrast, false positives from the GPT models seem to stem from a less conservative approach in determining comorbidity presence based on inferred context. These cases are also more likely to produce less reproducible responses due to the nondeterministic nature of LLMs. In these instances, physicians adopted a more conservative criterion to establish the ground truth, considering an unreported comorbidity only when the medication or context necessarily implied it. Whether this conservative approach is preferable to the criteria used by GPT models requires an analysis of complete medical histories to confirm or refute the presence of the comorbidity.

Discrepancies arising from variations in criteria interpretation could be mitigated by using prompts with clearer instructions on interpreting different comorbidities. This underscores the importance of refining prompts to enhance the consistency and accuracy of LLM-generated responses in clinical contexts.

Despite the remarkable capacity of current LLMs as potential tools for data mining in clinical reports, questions arise regarding the practical utility of this real-world data for research and the generation of real-world evidence [23]. The variability, subjectivity, and lack of structure in these reports can compromise the quality and reliability of extracted data, affecting its applicability in clinical research contexts. Therefore, while LLMs represent a promising innovation to address the limitations of unstructured data, implementing more structured clinical recording practices could provide a more sustainable and reliable solution for generating real-world clinical evidence. This duality emphasizes the need for a balanced approach that integrates advanced AI technology with robust clinical data management practices.

Future research should concentrate on refining prompt design and expanding the applications of LLMs across various medical fields. Additionally, exploring the performance of new open-source LLMs that can be run locally is essential, as this approach helps to avoid data protection and privacy issues associated with transmitting clinical data outside of the local infrastructure.

## Conclusion

This study has shown that, with carefully designed prompts, the OpenAI LLMs examined demonstrate competence comparable to, and in some cases superior to, that of medical specialists in interpreting and extracting relevant information from clinical reports, even when dealing with complex and ambiguously written texts. Considering their superior efficiency in terms of time and costs, along with their seamless integration with databases and other applications, these models emerge as a preferable option for data mining and structuring information in large collections of clinical reports. This highlights the potential of LLMs to enhance real-world data utilization by efficiently extracting structured information from extensive volumes of clinical texts, which is crucial for generating high-quality real-world evidence. Nonetheless, continuous evaluation of these models is essential to enhance their accuracy and applicability, while also emphasizing the importance of advancing towards more structured clinical records.

## Authors and collaborators

- **Study Idea and Design**: Amadeo Wals Zurita and Héctor Miras del Rio
- **Data Collection**: Carlos Míguez Sánchez, David Muñoz Carmona, María Rubio Jiménez, Nerea Ugarte Ruiz de Aguirre, Cristina Nebrera Navarro.
- **Data Analysis**: Héctor Miras del Rio and Amadeo Wals Zurita
- **Results Interpretation**: Héctor Miras del Rio and Amadeo Wals Zurita
- **Manuscript Writing**: Héctor Miras del Rio and Amadeo Wals Zurita
- **Critical Review and Editing**: all authors
- **Final Approval**: all authors
- **Data Anonymization and Randomization**: Data was pseudonymized by Andalusian Health Service technicians according to the GDPR regulation ensuring the technical and functional separation between the research team and those who perform the pseudonymization.

## Conflict of Interest

The authors of this scientific work declare that there are no conflicts of interest that could influence the results or conclusions of the conducted research. Furthermore, we emphasize that this study has not received any external funding. Our research was conducted independently, with the primary purpose of contributing to the advancement of knowledge in the corresponding area.

## Data Availability

All data produced in the present study are available upon reasonable request to the authors

https://github.com/RFMacarena/openaiAPIscript_forsharing.

## Abbreviations

cNLP: clinical natural language processing
EHR: electronic health record
LLM: large language model
JSON: JavaScript Object Notation
API: application programming interface
GPT: generative pre-trained transformers
RWD: real world data

## Notes

### Competing Interest Statement

The authors have declared no competing interest.

### Funding Statement

This study did not receive any funding

### Author Declarations

On 18-1-2024, a favorable opinion was issued by the Research Ethics Committee of the Virgen Macarena and Virgen del Rocio University Hospitals. EC_IA_V1 (Version 1-Dec-2023).

### Summary of Updates

The confusing or redundant phrases have been removed. Concepts have been added and clarified. Errors in the table and figure captions have been corrected. We have also added some bibliographic references to support some of our statements in the paper.

